# SYSTEMATIC REVIEW PROTOCOL: The reporting standards of Randomised Clinical Trials in leading medical journals between 2019-2020

**DOI:** 10.1101/2020.07.06.20147074

**Authors:** Mairead McErlean, Jack Samways, Peter Godolphin, Yang Chen

## Abstract

This is a systematic review protocol which outlines the basis and methodology for our intended review which at the time of writing is in the study screening phase. Our aim is to answer the fundamental questions:

To systematically identify RCTs published in the four leading medical journals between January 1st 2019 to May 31st 2020.

To assess the quality of reporting of such RCTs using the CONSORT 2010 statement.

To identify any association with medical specialty or size or type of RCT and the rate of adherence to the CONSORT 2010 statement.

## BACKGROUND

Randomised controlled trials (RCTs) remain the gold standard study design used to evaluate the safety and effectiveness of healthcare interventions.[1] When published in academic journals, years worth of work which cost hundreds of millions of dollars may be summarised in fewer than 3500 words.

It is imperative that the reporting quality of an RCT is transparent and clear to allow readers from the academic and scientific community to analyse and understand the study design and results which may change clinical practice. In an attempt to improve trial reporting, the Consolidated Standards of Reporting Trials (CONSORT) statement was developed in 1996.[2] Since its inception there have been significant updates in 2001 and 2010[3,4].

The current iteration is almost 10 years old and consists of a 25-item checklist for correct trial reporting. Studies have shown poor reporting standards in RCTs, particularly so in areas concerning trial methodology. [5-7] More recently in a field such as Cardiology with many RCTs a systematic review of reporting standards published in 2016 highlighted there is still room for improvement. Using the specific area of Heart Failure with preserved ejection fraction as an exemplar, of RCTs published after CONSORT 2010, the mean CONSORT score was only 63.8% (SD 18.1%). [8]

We wish to examine what the standard is now, in the highest impact general medical journals within academic medicine: The New England Journal of Medicine (NEJM), The Lancet, The Journal of the American Medical Association (JAMA) and The British Medical Journal (BMJ). Our assumption is that as prestigious journals who have all signed up to the CONSORT agreement, 25 years after its inception, we anticipate a high standard of adherence.

### Objectives

This review seeks to answer the fundamental questions:

- To systematically identify RCTs published in the four leading medical journals between January 1^st^ 2019 – May 31^st^ 2020
- To assess the quality of reporting of such RCTs using the CONSORT 2010 statement
- To identify any association with medical specialty or size or type of RCT and the rate of adherence to the CONSORT 2010 statement.

## METHODS

This protocol has been prepared in accordance with the guidance issued in the Preferred Reporting Items for Systematic Review and Meta-Analysis Protocols (PRISMA-P) 2015 statement.^9^

We will follow the Preferred Reporting Items for Systematic Reviews and Meta-Analyses (PRISMA) statement and Methodological Expectations of Cochrane Intervention Reviews (MECIR) standards for reporting systematic reviews.^10,11^

Once finalised, this protocol will be registered on the PROSPERO database and the registration number will be listed here. Post-registration changes to the protocol, if necessary, will be detailed under the PROSPERO record with the accompanying rationale.

## ELIGIBILITY CRITERIA

### Studies

- Randomised clinical trials in humans
- Between January 1^st^ 2020 – May 31^st^ 2020
- Only English language publications will be included
- Published in the New England Journal of Medicine (NEJM), The Lancet, The Journal of the Medical Association (JAMA) and The British Medical Journal (BMJ).
- Not a follow up analysis of a previously published trial result

### Participants

- Patients included in randomised clinical trials

### Interventions

- Medicinal product or other study intervention, including medical procedure or device

### Comparator

- Active control (standard of care) or placebo or other control

### Outcomes

- Adherence to CONSORT 2010 statement

### Data collection

- Area of medicine RCT is related to e.g. cardiovascular, oncology etc.
- Design of RCT – e.g. multi-arm adaptive, pilot or feasibility trial etc.
- Sample size
- Single centre or Multicentre
- Funding source

### Search strategy

To identify RCTs for inclusion we will filter our search to include all articles from NEJM, JAMA, Lancet and BMJ. A time filter will be applied to obtain results relevant to the study period. The Cochrane Highly Sensitive Search Strategies for identifying randomized trials filter will be used to identify only RCTs.^12^

An additional check with an expanded search filter used by the University of Texas will also be employed.^13^ The detailed search strategy is given in Appendix 1.

*Electronic searches: MEDLINE only* [*given that the target journals of our search are all indexed on this*].

### Study selection

Records arising from the literature search will be stored in a citation library using the software program Zotero. Duplicates will be removed from the results. Two reviewers (MM & JS) will screen titles and remove any reports that are clearly not eligible. These reviewers will then independently screen abstracts for potentially eligible studies. Full text reports will be retrieved for studies identified by at least one reviewer. Disagreements will be resolved by consensus and discussion with a third author (YC).

### Data extraction

Data will be extracted from study reports independently and in duplicate by reviewers for each eligible study. Each RCTs reporting quality will be scored against the CONSORT 2010 Statement.

### Data synthesis

We will not carry out any meta-analyses as the aim of this study is to assess the reporting of RCTs rather than examining their clinical outcomes. A systematic qualitative synthesis will be provided of the CONSORT scores and relevant trial characteristics of the included studies. Mean adherence to CONSORT for RCTs categorised by parent medical specialty, design and size will be reported.

### Publication-bias

We are not planning to specifically assess publication bias.

## Data Availability

The protocol is posted in full.

## AUTHOR CONTRIBUTIONS

MM & YC conceived the study. YC created the search strategy. All authors contributed to critical revision of the protocol for important intellectual content and approved the final version. YC is the guarantor.

## APPENDIX 1 SEARCH STRATEGY

### Medline (OvidSP)

1. JAMA.mp. or journal of the american medical association.jn. [mp=title, abstract, heading word, drug trade name, original title, device manufacturer, drug manufacturer, device trade name, keyword, floating subheading word, candidate term word]
2. bmj.mp. or british medical journal.jn. [mp=title, abstract, heading word, drug trade name, original title, device manufacturer, drug manufacturer, device trade name, keyword, floating subheading word, candidate term word]
3. lancet.jn.
4. nejm.mp. or new england journal of medicine.jn. [mp=title, abstract, heading word, drug trade name, original title, device manufacturer, drug manufacturer, device trade name, keyword, floating subheading word, candidate term word]
5. 1 or 2 or 3 or 4
6. (“clinical trial” or “clinical trial, phase i” or “clinical trial, phase ii” or clinical trial, phase iii or clinical trial, phase iv or controlled clinical trial or “multicenter study” or “randomized controlled trial”).pt. or double-blind method/ or clinical trials as topic/ or clinical trials, phase i as topic/ or clinical trials, phase ii as topic/ or clinical trials, phase iii as topic/ or clinical trials, phase iv as topic/ or controlled clinical trials as topic/ or randomized controlled trials as topic/ or early termination of clinical trials as topic/ or multicenter studies as topic/ or ((randomi?ed adj7 trial*) or (controlled adj3 trial*) or (clinical adj2 trial*) or ((single or doubl* or tripl* or treb*) and (blind* or mask*))).ti,ab,kw. or (“4 arm” or “four arm”).ti,ab,kw.
7. 5 and 6
8. limit 7 to yr=“2019 -Current”

